# Participatory Approaches in Community Health in light of the COVID-19 Pandemic: A Scoping Review Protocol

**DOI:** 10.1101/2023.08.24.23294551

**Authors:** Annika Frahsa, Harvy Joy Liwanag, Cristopher Kobler, Aziz Mert Ipekci, Beatrice Minder, Diana Schow

## Abstract

**Background:** Participatory approaches are considered essential to ensure community health in the context of the COVID-19 pandemic. Previous reviews on community participation have explored different aspects of participation in specific contexts, such as public health emergencies, but none has examined participatory approaches both in depth and in breadth across diverse activities during the COVID-19 pandemic and considering diverse communities in all country contexts. This scoping review seeks to: (a) provide an overview of participatory approaches in terms of the features and depth of participation, the breadth of the communities and stakeholders involved, and for what types of activities and interventions in light of the COVID-19 pandemic across all country contexts; (b) explore the challenges and facilitators of participation processes; and (c) analyse to what extent participation impacts community health, including health equity, in the context of a public health emergency.

**Methods:** We developed this protocol following the latest JBI guidance on scoping reviews. A comprehensive search strategy combining the concepts of participation, community health, and COVID-19 was used to search the databases of Medline/Ovid, Embase.com, Cochrane CENTRAL, Web of Science, APA PsycInfo/Ovid, Global Health/Ovid, ERIC/OvidSP, CINAHL/EBSCOhost, ClinTrials.gov, and the grey literature through Google Scholar. At least two reviewers will perform screening of titles/abstracts and full text using the inclusion and exclusion criteria defined in this protocol. Article characteristics and data on participatory approaches and community health will be charted to provide an overview of the literature, map the variations in participatory approaches and community health, and explore patterns in the links between participation, community health, and the type of activities to address the challenges related to the COVID-19 pandemic.

**Discussion:** We anticipate that review findings will contribute to advance innovative thinking about community participation and facilitating better application and integration of participatory approaches to ensure community health in a future public health emergency or in building back better fairer in the new normal.

## INTRODUCTION

The COVID-19 pandemic has exacerbated inequities and severely impacted health and wellbeing in many contexts.^1^ To respond effectively to public health emergencies, participatory approaches are essential for community health.^2^ In the early phase of the pandemic, community participation was already viewed as important to reach marginalised populations and support equity-informed responses.^3^ Most recently in the context of “building back fairer”, participation is seen to contribute to creating community resilience^4^ and promoting pandemic preparedness.^5,6^ Outside the setting of pandemics, community participation has been emphasised both in the Alma Ata Declaration^7^ and the World Health Organization (WHO) Ottawa Charter for Health Promotion^8^, and in different national programmes and foundational documents, such as the constitution of National Health Service England^9^, the principles behind the German cooperation for equity in health,^10^ and landmark legislation to strengthen the health sector in low- and middle-income countries (LMICs) like South Africa^11^ and Brazil.^12^

Participatory approaches in community health have often been presented as a continuum, ranging from dissemination and outreach *to communities* on one end of the spectrum, to co-decision-making and shared leadership *with communities* on the other end of the spectrum.^13^ Others have presented participatory approaches via the dichotomy of two main perspectives: (a) the emancipatory or empowerment argument and (b) the usefulness or utilitarian argument.^14,15^ The *emancipatory* or *empowerment argument* views participation as a means to address inequities, reduce marginalisation, and rebalance power relations.^16,17^ This perspective on participation is widely represented in community-based participatory research for health (CBPR), an approach Wallerstein *et al.* defined as “collaborative efforts among community, academic, and other stakeholders who gather and use research and data to build on the strengths and priorities of the community for multilevel strategies to improve health and social equity.”^18^ Within this approach, research begins with a topic of interest to a specific community and members are involved as partners in iterative cycles of action and reflection for change and are trained as co-researchers. Given its equity focus, CBPR addresses health issues as much as community assets, resources, and capacities, and empowerment.

In the *usefulness* or *utilitarian argument,* participation is viewed as a strategy to promote the efficient or effective implementation of health policies or programmes.^14^ Within this perspective, research and action tends to start with a topic of concern to researchers, such as the need to recruit specific population subgroups or communities into clinical trials or surveys, or to reach out to diverse communities with a given intervention study.^19^ Community members are involved to (re-)contextualize expert-driven scientific planning or findings and the focus tends to be on individual health or behaviour-related outcomes with little consideration of power relations or community capacity-building. These two central yet different perspectives on community participation are also reflected in the discourse on participatory approaches in community health in the context of the COVID-19 pandemic.

### What we know about participation and community health

Initial reviews on participatory approaches in community health linked to the COVID-19 pandemic have been published in the literature, with many of these reviews focussing on certain aspects of participation or in specific contexts. For example, one scoping review examined “community engagement and involvement” and mapped the stakeholders particularly in urban poor settings in LMICs.^20^ There is also a systematic review still in progress that aims to examine “community engagement” with a global scope but specifically on how participation supports COVID-19 vaccine uptake.^21^ One rapid evidence review explored “community engagement” approaches during infectious disease outbreaks prior to COVID-19 (i.e., Ebola, Zika, SARS, MERS, and H1N1 outbreaks) and identified community engagement actors and their functions.^3^

Outside the context of public health emergencies, several reviews on community participation have been published, starting with the first reviews on CBPR by Israel *et al.* in 1998^22^ and by Salimi *et al.* in 2012.^23^ A scoping review that specifically explored the literature on “community-engaged research” has identified up 100 reviews on the topic between 2005-2018.^24^ One systematic review in 2019 on “community participation” in health services focused on high-income countries (HICs) and concluded that participation had a positive impact when substantiated by strong organisational and community processes,^25^ while an earlier systematic review in 2015 assessed the extent, nature, and quality of “community participation” in health systems research on LMICs and concluded that participation was variable.^16^

### Knowledge gaps this review will address

This scoping review will contribute to the body of knowledge on participation and community health by addressing three key gaps that remain. *First,* there remains a lack of a comprehensive analysis of participatory approaches and their variations as represented in the literature. Such an analysis should not only describe participatory approaches as wide-ranging but also systematically characterise the depth of participation across a spectrum of low-to-high participation, including which stakeholders participate in the process and the balance of power shared between them. An analysis of participatory approaches is comprehensive when it also examines the nuances in the narratives of participation given the differences in the terms used to refer to participation. The *second* gap has to do with the need for a synthesis of the “complete picture” of the breadth of participatory approaches in the context of the COVID-19 pandemic involving all types of communities, covering all categories of activities and interventions, and across all geographic and country income contexts. *Third,* it remains unclear how exactly participation has impacted community health in the setting of the COVID-19 pandemic. The wealth of information from numerous COVID-19-related publications that have been added to the literature in recent years offers a strategic opportunity for this scoping review to address these gaps.

### Review objective and questions

To achieve a comprehensive, extensive, and systematic understanding of participatory approaches in community health in light of the COVID-19 pandemic, this scoping review aims to provide an overview of participatory approaches in terms of the features and depth of participation, the breadth of the communities and stakeholders involved, and for what types of activities and interventions in light of the pandemic across all country contexts. We will also explore the challenges and facilitators of the participation process and how and to what extent participation impacts community health, including health equity, in the setting of a public health emergency. Our three main review questions are as follows:

i. What were the participatory approaches applied in community health in light of the COVID-19 pandemic according to depth of participation, communities involved, type of pandemic response, and country contexts?
ii. What were the main challenges and facilitators for participatory approaches in community health in the COVID-19 pandemic?
iii. How did participatory approaches impact community health, including health equity, in the context of the COVID-19 pandemic?

To the best of our knowledge, this is the first scoping review of participatory approaches with an extensive coverage of all types of participation, community groups, categories of interventions in the context of COVID-19, and across all country contexts. It is important to highlight that, in addressing the *third* review question regarding impacts, our goal here is not to demonstrate a causal link between participation and community health and particularly not on individual-level outcomes. Instead, we are interested in a deeper understanding of participation as a process,^26^ including a reflective analysis of its relevant domains that support concrete interventions to respond to the COVID-19 pandemic. We will explore the possibility of developing a preliminary theory of change or logic model^27^ that could explain why and how participation contributes to community health in the setting of the COVID-19 pandemic. We anticipate that our findings will contribute not only in addressing the knowledge gaps enumerated above but also in advancing innovative thinking about community participation and facilitating better application and integration of participatory approaches to ensure community health in a future public health emergency^28^ or in building back better fairer in the new normal.^1,4^

## METHODS

We developed this protocol following the latest guidance for scoping review protocols by Peters *et al.*^29^ The methods of a scoping review is suitable when the goal is to provide an overview of the literature on a topic of interest. We searched the Open Science Framework (OSF)^30^ and PROSPERO registry^31^ and did not find a similar review currently in progress at the time of this writing. This scoping review protocol was registered on OSF on 24 Aug 2023.

### PCC Framework

We referred to the Population-Concept-Context (PCC) framework^29^ to develop our search strategy. Our *Population* of interest is the community in “community health.” Within the field of health promotion, community health refers to the health status of a group of people, defined by their geographical proximity, shared values, culture, norms or special interests and the actions and conditions to promote, protect and preserve their health.^32^ Typically, a community is arranged in a social structure based on relationships that have been developed over a period of time. Its members have some awareness of their group identity and a commitment to meet their shared needs. Community health is not the same as “population health” that is often used in biomedical or epidemiological research which only refers to the health status of people generally without defining them as particular groups. Our *Concept* of interest is “participation” which could be understood in a spectrum of high to low as illustrated in Arnstein’s ladder of citizen participation: from providing information, to consultation, to involvement through regular interactions, to collaboration that require shared decision-making, to community self-organisation.^33^ There are several overlapping terms that are also used but may refer to the same idea of participation such as participatory action research, action research, participatory research, youth participatory action research, public involvement, practitioner research, collaborative research, citizen science, street science, participatory health research,^15^ as well as co-creation, collaboration, co-design, engagement.^34^ Our search strategy and planned analysis will cover all variations in the use of these terms that refer to participation. Finally, our *Context* of interest will be the COVID-19 pandemic in any country regardless of geographic location or income classification.

### Eligibility criteria

We will include articles that fulfil the following criteria:

- The article is about the COVID-19 pandemic;
- The article describes a participatory approach implemented in the context of the COVID-19 pandemic;
- The participatory approach in the article involves a particular community of people. All of the above elements must be present for an article to be considered eligible.

We will exclude articles if:

- The article is in the category of “opinion-based” articles such as, but not limited to, commentaries, perspectives, letters to the editor, viewpoints, correspondence, editorials, news feature, etc.;
- The participatory approach in the article is for addressing a non-health challenge (e.g., loss of income, lack of transportation, etc.), unless the challenge is discussed in relation to health;
- The community/ies referred to in the article are in a healthcare facility setting or in a biomedical/clinical context; or refer to specific patient groups seeking treatment for a particular service (e.g., prenatal care), community of health professionals or trainees (e.g., community pharmacists, medical students, etc.), or virtual communities (e.g., online communities, social media communities, etc.).

Our process for including/excluding an article during screening is summarised by the following flowchart:

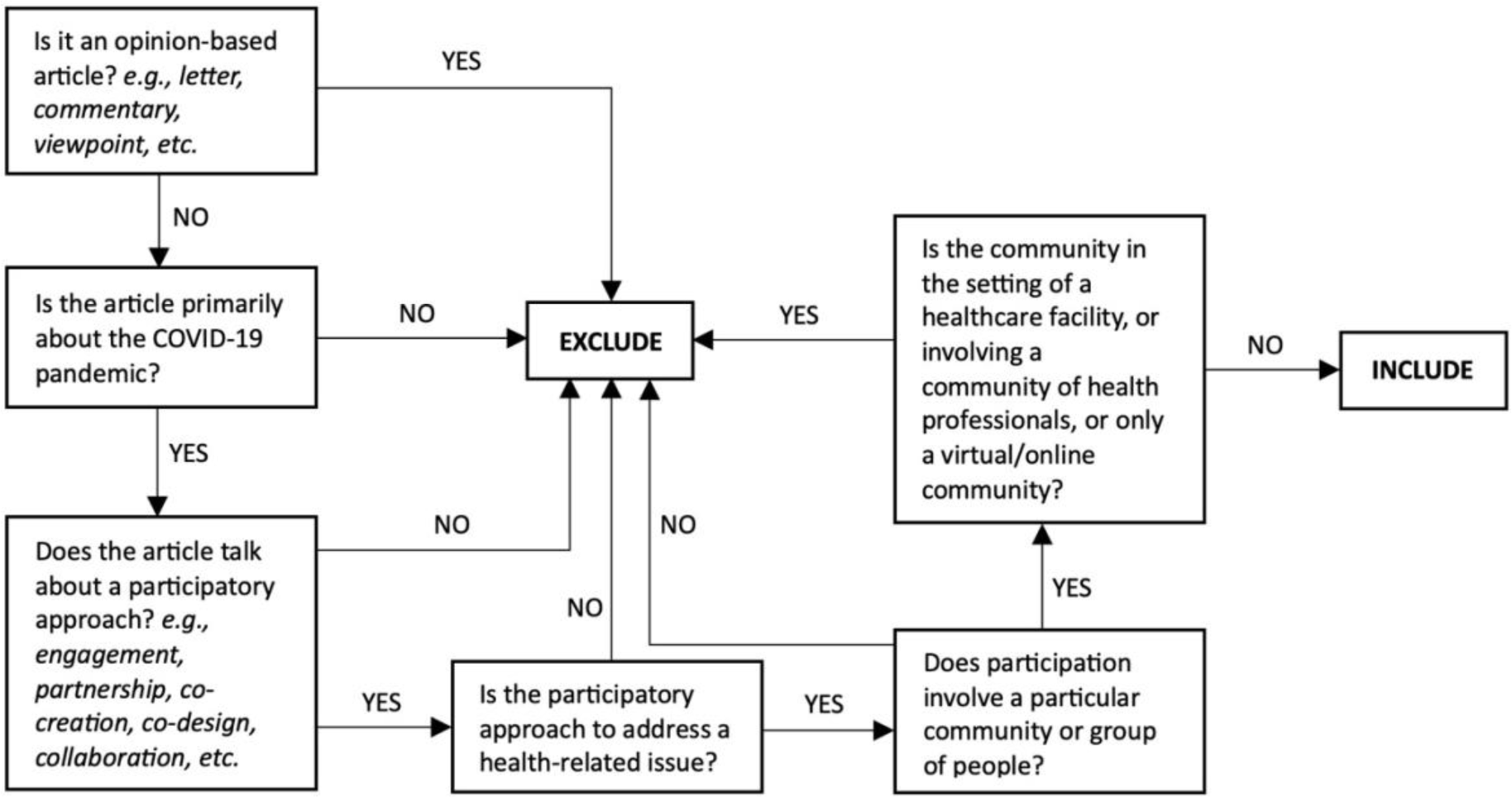

We will apply no date restriction, although we do not anticipate screening any article before 2020, or the year when COVID-19 became a pandemic, because our search strategy specifically searched for COVID-19-related publications. We will also apply no language restrictions. Our team includes reviewers who are fluent in English, German, and Spanish. During title/abstract screening, we will use DeepL Translate (DeepL SE, Cologne, Germany) to translate non-English articles and assess if the article would be considered further. A non-English article included in full text screening will be translated by consulting colleagues who are fluent in the language of the article.

### Information sources

Nine bibliographic databases were searched until June 2nd 2023: Medline/Ovid, Embase.com, Cochrane CENTRAL, Web of Science, APA PsycInfo/Ovid, Global Health/Ovid, ERIC/OvidSP, CINAHL/EBSCOhost, ClinTrials.gov. Additional articles (including grey literature) were retrieved based on the first 200 references from Google Scholar. Duplicate records were removed using a fully automated deduplication solution, Deduklick.^35^ Since we are not excluding review articles, we will also review their list of references and assess the eligibility of each reference.

### Search strategy

The complete search strategy is presented in the Appendix.

### Records management and screening process

Screening and management of records will be performed using Endnote^TM^ (Clarivate, PA, USA) with a duplicate set of records maintained using Zotero (CDS, VA, USA) as back-up. Records will be stored securely using the OneDrive (M365) (Microsoft, WA, USA) campus account of the University of Bern. Both the phases of title/abstract screening and full text screening will involve two reviewers working independently. Any conflicts in the inclusion/exclusion of an article will be resolved by discussion between the two reviewers or by consulting the project leader (AF). We will summarise the screening process through a PRISMA flow diagram in the final manuscript. Charting of data will also be performed by two reviewers using Microsoft Excel® (Microsoft, WA, USA) and the information collected will be reconciled through discussion. We will not be performing quality appraisal to assess bias in articles following the guidance for scoping reviews^29^ because our goal is to map the body of literature and not to answer a focused review question on effectiveness.

### Data items to be charted

i. Characteristics of included articles

- Authors and year of publication
- Country of the main affiliation of the first author
- Country/ies where the study was undertaken

- Further grouped according to country income level based on the World Bank classification^36^ and according to the regions of the World Health Organization^37^
- Type of article (e.g., research article) and study design (e.g., mixed methods)
- Main discipline/s of the study (e.g., economics, political science, etc.)
- At which phase of the pandemic (e.g., early phase, during, or post-pandemic)
ii. Participatory approaches for community health

### Description of participation

- Term/s used to refer to participation (e.g., co-creation, engagement, etc.)
- How participation was defined in the article
- Narrative or basis for participation (e.g., emancipatory, utilitarian, or another argument)
- Type of activity or health intervention/s that involved participation, whether to address COVID-19 directly (e.g., COVID-19 vaccination) or indirectly (e.g., support for loneliness)
- How participation was implemented

- Brief description of the process
- During which stage of the public health action cycle^38^ (e.g., agenda setting, problem definitions, planning, priority setting, implementation, monitoring/evaluation, quality management, advocacy)
- Depth of participation (low-to-high, referring to Arnstein’s ladder of participation^33^)
- Challenges and facilitators to participation

### Description of community health

- Definition of community health (if any given)
- Community groups involved in the participation according to

- Gender (e.g., women’s group)
- Socioeconomic class (e.g., middle-income families)
- Age (e.g., teenagers, elderly)
- Livelihood (e.g., farming communities)
- Race, ethnicity, or nationality (e.g., Asian)
- Religious beliefs (e.g., Muslim community)
- Vulnerable groups (e.g., migrants, refugees)
- Other group categories
- Specific individual actors in the community who were involved, including which actors possessed more (or less) power in influencing the process
- Outcomes at the community level that were targeted by the participatory approach

- Health equity outcomes
- Other outcomes related to community health and wellbeing

### Planned data synthesis

We will use descriptive statistics to present an overview of the characteristics of the included studies through tables and graphs. We will perform content analysis^39^ in examining variations in participatory approaches and community health and use mapping^40^ to visualise patterns in the links between participation, community health, and the type of intervention to address the COVID-19 pandemic. We will explore the possibility of developing a theoretical framework or logic model^27^ to illustrate how and why participation impacts community health, including how it contributes to health equity, in a setting of a public health emergency. Finally, we will identify new research questions that will build on our review findings and propose a research agenda and/or a subsequent study to test our theory empirically.

Although this is an *a priori* protocol, we also recognise that the presentation of findings in the final manuscript could evolve because of the iterative process of data charting and synthesis.^29^ We will report deviations from this protocol in the final manuscript.

## Data Availability

This is a protocol for a scoping review.

## Contributions

AF conceptualised the review and is the guarantor of the project. AF and HJL developed the review protocol with inputs from BM. BM designed the search strategies and performed the literature searches. HJL will perform management of records. AF, HJL, CK, MIA, and DS will conduct article screening. AF, HJL, and CK will perform data charting and analysis. All authors approved this protocol and will contribute to writing the final manuscript.

## APPENDIX

### Documentation of search strategies

Combination of concepts: 1) AND 2) AND 3) + adding filters 4)

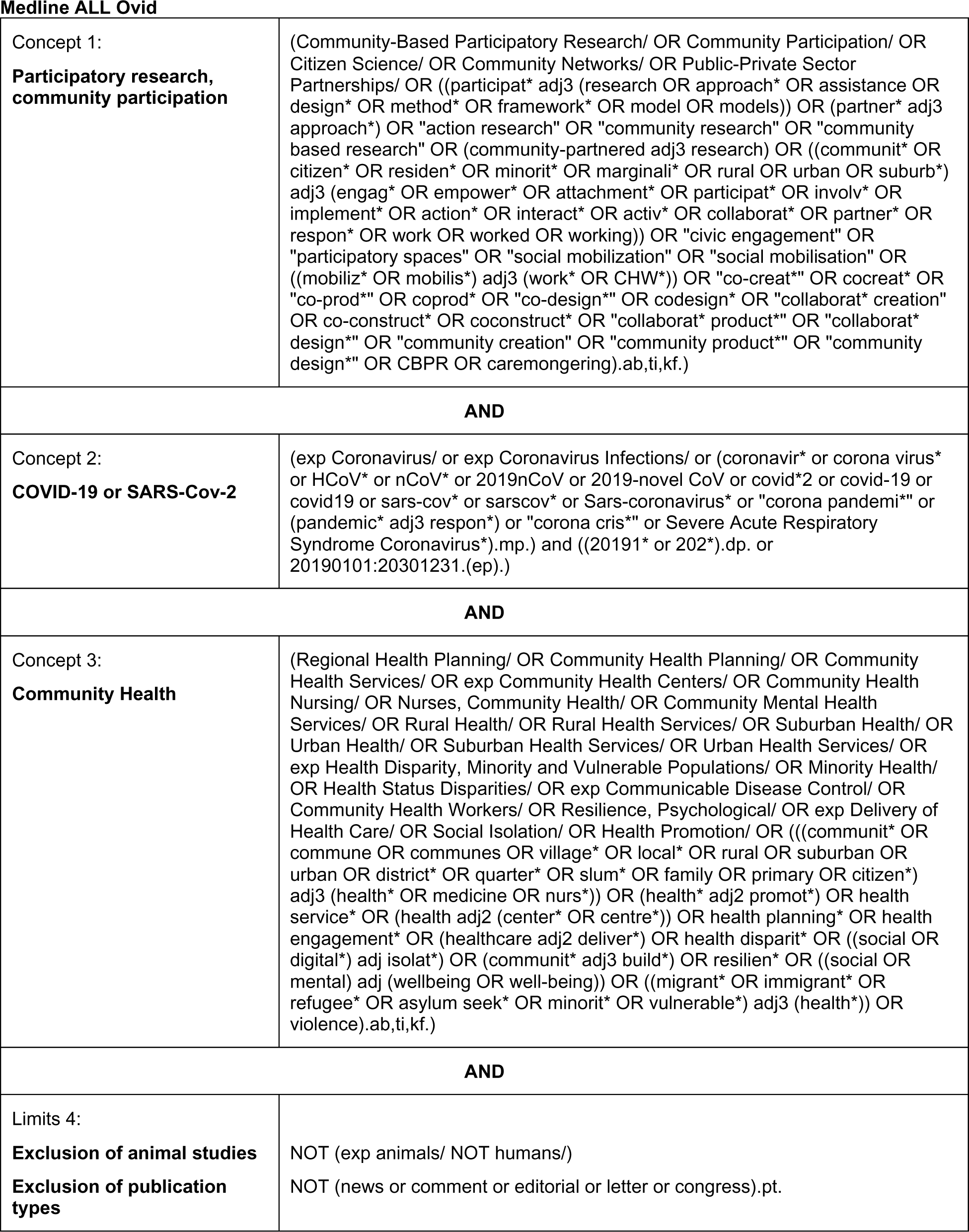

#### One paragraph search strategies

##### Medline ALL Ovid

(Community-Based Participatory Research/ OR Community Participation/ OR Citizen Science/ OR Community Networks/ OR Public-Private Sector Partnerships/ OR ((participat* adj3 (research OR approach* OR assistance OR design* OR method* OR framework* OR model OR models)) OR (partner* adj3 approach*) OR “action research” OR “community research” OR “community based research” OR (community-partnered adj3 research) OR ((communit* OR citizen* OR residen* OR minorit* OR marginali* OR rural OR urban OR suburb*) adj3 (engag* OR empower* OR attachment* OR participat* OR involv* OR implement* OR action* OR interact* OR activ* OR collaborat* OR partner* OR respon* OR work OR worked OR working)) OR “civic engagement” OR “participatory spaces” OR “social mobilization” OR “social mobilisation” OR ((mobiliz* OR mobilis*) adj3 (work* OR CHW*)) OR “co-creat*” OR cocreat* OR “co-prod*” OR coprod* OR “co-design*” OR codesign* OR “collaborat* creation” OR co-construct* OR coconstruct* OR “collaborat* product*” OR “collaborat* design*” OR “community creation” OR “community product*” OR “community design*” OR CBPR OR caremongering).ab,ti,kf.) AND (exp Coronavirus/ or exp Coronavirus Infections/ or (coronavir* or corona virus* or HCoV* or nCoV* or 2019nCoV or 2019-novel CoV or covid*2 or covid-19 or covid19 or sars-cov* or sarscov* or Sars-coronavirus* or “corona pandemi*” or (pandemic* adj3 respon*) or “corona cris*” or Severe Acute Respiratory Syndrome Coronavirus*).mp.) and ((20191* or 202*).dp. or 20190101:20301231.(ep).) AND (Regional Health Planning/ OR Community Health Planning/ OR Community Health Services/ OR exp Community Health Centers/ OR Community Health Nursing/ OR Nurses, Community Health/ OR Community Mental Health Services/ OR Rural Health/ OR Rural Health Services/ OR Suburban Health/ OR Urban Health/ OR Suburban Health Services/ OR Urban Health Services/ OR exp Health Disparity, Minority and Vulnerable Populations/ OR Minority Health/ OR Health Status Disparities/ OR exp Communicable Disease Control/ OR Community Health Workers/ OR Resilience, Psychological/ OR exp Delivery of Health Care/ OR Social Isolation/ OR Health Promotion/ OR (((communit* OR commune OR communes OR village* OR local* OR rural OR suburban OR urban OR district* OR quarter* OR slum* OR family OR primary OR citizen*) adj3 (health* OR medicine OR nurs*)) OR (health* adj2 promot*) OR (health* adj2 service*) OR (health adj2 (center* OR centre*)) OR health planning* OR health engagement* OR (healthcare adj2 deliver*) OR health disparit* OR ((social OR digital*) adj isolat*) OR (communit* adj3 build*) OR resilien* OR ((social OR mental) adj (wellbeing OR well-being)) OR ((migrant* OR immigrant* OR refugee* OR asylum seek* OR minorit* OR vulnerable*) adj3 (health*)) OR violence).ab,ti,kf.) NOT (exp animals/ NOT humans/) NOT (news or comment or editorial or letter or congress).pt.

##### Embase.com

(’participatory research’/exp OR ‘participatory management’/de OR ‘community participation’/exp OR ‘citizen science’/de OR ‘action research’/exp OR ‘civic engagement’/de OR ‘public-private partnership’/de OR ‘social network’/de OR ((participat* NEAR/3 (research OR approach* OR assistance OR design* OR method* OR framework* OR model OR models)) OR (partner* NEAR/3 approach*) OR ‘action research’ OR ‘community research’ OR ‘community based research’ OR (community-partnered NEAR/3 research) OR ((communit* OR citizen* OR residen* OR minorit* OR marginali* OR rural OR urban OR suburb*) NEAR/3 (engag* OR empower* OR attachment* OR participat* OR involv* OR implement* OR action* OR interact* OR activ* OR collaborat* OR partner* OR respon* OR work OR worked OR working)) OR ‘civic engagement’ OR ‘participatory spaces’ OR ‘social mobilization’ OR ‘social mobilisation’ OR ((mobiliz* OR mobilis*) NEAR/3 (work* OR CHW*)) OR ‘co-creat*’ OR cocreat* OR ‘co-prod*’ OR coprod* OR ‘co-design*’ OR codesign* OR ‘collaborat* creation’ OR co-construct* OR coconstruct* OR ‘collaborat* product*’ OR ‘collaborat* design*’ OR ‘community creation’ OR ‘community product*’ OR ‘community design*’ OR CBPR OR caremongering):ab,ti,kw) AND ((’coronavirus disease 2019’/exp OR ‘Coronavirus infection’/de OR ‘Severe acute respiratory syndrome coronavirus 2’/exp OR (coronavir* or ‘corona virus*’ or HCoV* or nCoV* or 2019nCoV or ‘2019-novel CoV’ or covid or covid-19 or covid19* or sars-cov* or sarscov* or Sars-coronavirus* or ‘corona pandemi*’ or (pandemic* NEAR/3 respon*) or ‘corona cris*’ or ‘Severe Acute Respiratory Syndrome Coronavirus*’):ab,ti,kw) AND [2019-3000]/py) AND (’community care’/exp OR ‘health care planning’/de OR ‘health care need’/exp OR ‘health care access’/exp OR ‘health center’/de OR ‘community health nursing’/de OR ‘mental health service’/exp OR ‘rural health’/de OR ‘rural health nursing’/de OR ‘rural health care’/de OR ‘urban health’/de OR ‘health services research’/de OR ‘vulnerable population’/exp OR ‘minority health’/de OR ‘health disparity’/de OR ‘health care disparity’/de OR ‘communicable disease control’/de OR ‘infection control’/de OR ‘quarantine’/exp OR ‘disease surveillance’/exp OR ‘health auxiliary’/de OR ‘community resilience’/de OR ‘psychological resilience’/de OR ‘health care delivery’/exp OR ‘health equity’/de OR ‘social isolation’/de OR ‘social stigma’/de OR ‘health promotion’/de OR ‘psychological well-being’/de OR (((communit* OR commune OR communes OR village* OR local* OR rural OR suburban OR urban OR district* OR quarter* OR slum* OR family OR primary OR citizen*) NEAR/3 (health* OR medicine OR nurs*)) OR (health* NEAR/2 promot*) OR ‘health service*’ OR (health NEAR/2 (center* OR centre*)) OR ‘health planning*’ OR ‘health engagement*’ OR (healthcare NEAR/2 deliver*) OR ‘health disparit*’ OR ((social OR digital*) NEAR/3 isolat*) OR (communit* NEAR/3 build*) OR resilien* OR ((social OR mental) NEAR/3 (wellbeing OR well-being)) OR ((migrant* OR immigrant* OR refugee* OR ‘asylum seek*’ OR minorit* OR vulnerable*) NEAR/3 (health*)) OR violence):ab,ti,kw) NOT ([animals]/lim NOT [humans]/lim) NOT ([Conference Abstract]/lim OR [Letter]/lim OR [Note]/lim OR [Editorial]/lim) NOT ‘preprint’/it

##### Cochrane CENTRAL

(((participat* NEAR/3 (research OR approach* OR assistance OR design* OR method* OR framework* OR model OR models)) OR (partner* NEAR/3 approach*) OR “action research” OR “community research” OR “community based research” OR (community-partnered NEAR/3 research) OR ((communit* OR citizen* OR residen* OR minorit* OR marginali* OR rural OR urban OR suburb*) NEAR/3 (engag* OR empower* OR attachment* OR participat* OR involv* OR implement* OR action* OR interact* OR activ* OR collaborat* OR partner* OR respon* OR work OR worked OR working)) OR “civic engagement” OR “participatory spaces” OR “social mobilization” OR “social mobilisation” OR ((mobiliz* OR mobilis*) NEAR/3 (work* OR CHW*)) OR (co NEXT creat*) OR cocreat* OR (co NEXT prod*) OR coprod* OR (co NEXT design*) OR codesign* OR (collaborat* NEXT creation) OR (co NEXT construct*) OR coconstruct* OR (collaborat* NEXT product*) OR (collaborat* NEXT design*) OR “community creation” OR (community NEXT product*) OR (community NEXT design*) OR CBPR OR caremongering):ab,ti,kw) AND ((coronavir* OR corona NEXT virus* OR corona NEXT pandemi* OR corona NEXT cris* OR (pandemic* NEAR/3 respon*) OR HCoV* OR nCoV* OR “2019 CoV” OR 2019nCoV* OR “2019-novel CoV*” OR covid OR covid19* OR sars-cov* OR sarscov* OR sars-coronavirus* OR sarscoronavir* OR Severe Acute Respiratory Syndrome Coronavirus*):ab,ti,kw) AND ((((communit* OR commune OR communes OR village* OR local* OR rural OR suburban OR urban OR district* OR quarter* OR slum* OR family OR primary OR citizen*) NEAR/3 (health* OR medicine OR nurs*)) OR (health* NEAR/2 promot*) OR health NEXT service* OR (health NEAR/2 (center* OR centre*)) OR health NEXT planning* OR health NEXT engagement* OR (healthcare NEAR/2 deliver*) OR health NEXT disparit* OR ((social OR digital*) NEAR/3 isolat*) OR (communit* NEAR/3 build*) OR resilien* OR ((social OR mental) NEAR/3 (wellbeing OR well-being)) OR ((migrant* OR immigrant* OR refugee* OR asylum-seek* OR minorit* OR vulnerable*) NEAR/3 (health*)) OR violence):ab,ti,kw)

##### Web of Science Core Collection* (via Clarivate)

TS=(((participat* NEAR/3 (research OR approach* OR assistance OR design* OR method* OR framework* OR model OR models)) OR (partner* NEAR/3 approach*) OR “action research” OR “community research” OR “community based research” OR (community-partnered NEAR/3 research) OR ((communit* OR citizen* OR residen* OR minorit* OR marginali* OR rural OR urban OR suburb*) NEAR/3 (engag* OR empower* OR attachment* OR participat* OR involv* OR implement* OR action* OR interact* OR activ* OR collaborat* OR partner* OR respon* OR work OR worked OR working)) OR “civic engagement” OR “participatory spaces” OR “social mobilization” OR “social mobilisation” OR ((mobiliz* OR mobilis*) NEAR/3 (work* OR CHW*)) OR co-creat* OR cocreat* OR co-prod* OR coprod* OR co-design* OR codesign* OR (collaborat* NEAR/1 creation) OR co-construct* OR coconstruct* OR (collaborat* NEAR/1 product*) OR (collaborat* NEAR/1 design*) OR “community creation” OR “community product*” OR “community design*” OR CBPR OR caremongering)) AND TS=((coronavir* OR “corona virus*” OR “corona pandemi*” OR “corona cris*” OR (pandemic* NEAR/2 respon*) OR HCoV* OR nCoV* OR “2019 CoV” OR 2019nCoV* OR “2019-novel CoV*” OR covid OR covid19* OR sars-cov* OR sarscov* OR sars-coronavirus* OR sarscoronavir* OR “Severe Acute Respiratory Syndrome Coronavirus*”)) AND TS=((((communit* OR commune OR communes OR village* OR local* OR rural OR suburban OR urban OR district* OR quarter* OR slum* OR family OR primary OR citizen*) NEAR/3 (health* OR medicine OR nurs*)) OR (health* NEAR/1 promot*) OR “health service*” OR (health NEAR/2 (center* OR centre*)) OR “health planning*” OR “health engagement*” OR (healthcare NEAR/2 deliver*) OR “health disparit*” OR (mitigat* NEAR/1 disparit*) OR ((social OR digital*) NEAR/3 isolat*) OR (communit* NEAR/3 build*) OR resilien* OR ((social OR mental) NEAR/3 (wellbeing OR well-being)) OR ((migrant* OR immigrant* OR refugee* OR asylum-seek* OR minorit* OR vulnerable* OR worker*) NEAR/3 (health*)) OR violence)) *Refined by: NOT Document Types: Proceedings Papers or Meeting Abstracts or Editorial Material or Letter* *Science Citation Index Expanded (1900-present) ; Social Sciences Citation Index (1900-present) ; Arts & Humanities Citation Index (1975-present) ; Conference Proceedings Citation Index-Science (1990-present) ; Conference Proceedings Citation Index-Social Science & Humanities (1990-present) ; Emerging Sources Citation Index (2018-present)

##### APA PsycINFO (via Ovid)

((Action Research/ or Community Involvement/ or ((participat* adj3 (research or approach* or assistance or design* or method* or framework* or model or models)) or (partner* adj3 approach*) or “action research” or “community research” or “community based research” or (community-partnered adj3 research) or ((communit* or citizen* or residen* or minorit* or marginali* or rural or urban or suburb*) adj3 (engag* or empower* or attachment* or participat* or involv* or implement* or action* or interact* or activ* or collaborat* or partner* or respon* or work or worked or working)) or “civic engagement” or “participatory spaces” or “social mobilization” or “social mobilisation” or ((mobiliz* or mobilis*) adj3 (work* or CHW*)) or “co-creat*” or cocreat* or “co-prod*” or coprod* or “co-design*” or codesign* or “collaborat* creation” or co-construct* or coconstruct* or “collaborat* product*” or “collaborat* design*” or “community creation” or “community product*” or “community design*” or CBPR or caremongering).ab,ti.) and (exp coronavirus/ or (coronavir* or corona virus* or HCoV* or nCoV* or 2019nCoV or 2019-novel CoV or covid*2 or covid-19 or covid19 or sars-cov* or sarscov* or Sars-coronavirus* or “corona pandemi*” or (pandemic* adj3 respon*) or “corona cris*” or Severe Acute Respiratory Syndrome Coronavirus*).mp.) and (exp community health/ or exp community services/ or community mental health centers/ or rural health/ or urban health/ or health care services/ or health disparities/ or “resilience (psychological)”/ or psychological endurance/ or exp health care delivery/ or social isolation/ or health promotion/ or (((communit* or commune or communes or village* or local* or rural or suburban or urban or district* or quarter* or slum* or family or primary or citizen*) adj3 (health* or medicine or nurs*)) or (health* adj2 promot*) or (health* adj2 service*) or (health adj2 (center* or centre*)) or health planning* or health engagement* or (healthcare adj2 deliver*) or health disparit* or ((social or digital*) adj isolat*) or (communit* adj3 build*) or resilien* or ((social or mental) adj (wellbeing or well-being)) or ((migrant* or immigrant* or refugee* or asylum seek* or minorit* or vulnerable*) adj3 health*) or violence).ab,ti.)) not (exp animals/ not humans/) not (letter or comment or editorial or abstract).dt.

##### Global Health (via Ovid)

((Participation/ or Public Participation/ or Social Participation/ or Community Involvement/ or Community Action/ or ((participat* adj3 (research or approach* or assistance or design* or method* or framework* or model or models)) or (partner* adj3 approach*) or “action research” or “community research” or “community based research” or (community-partnered adj3 research) or ((communit* or citizen* or residen* or minorit* or marginali* or rural or urban or suburb*) adj3 (engag* or empower* or attachment* or participat* or involv* or implement* or action* or interact* or activ* or collaborat* or partner* or respon* or work or worked or working)) or “civic engagement” or “participatory spaces” or “social mobilization” or “social mobilisation” or ((mobiliz* or mobilis*) adj3 (work* or CHW*)) or “co-creat*” or cocreat* or “co-prod*” or coprod* or “co-design*” or codesign* or “collaborat* creation” or co-construct* or coconstruct* or “collaborat* product*” or “collaborat* design*” or “community creation” or “community product*” or “community design*” or CBPR or caremongering).ab,ti,id.) and (exp severe acute respiratory syndrome coronavirus 2/ or (coronavir* or corona virus* or HCoV* or nCoV* or 2019nCoV or 2019-novel CoV or covid*2 or covid-19 or covid19 or sars-cov* or sarscov* or Sars-coronavirus* or “corona pandemi*” or (pandemic* adj3 respon*) or “corona cris*” or Severe Acute Respiratory Syndrome Coronavirus*).mp.) and (exp community health/ or exp community health services/ or public services/ or mental health/ or rural health/ or health inequalities/ or exp disease control/ or exp community health workers/ or endurance/ or social isolation/ or health promotion/ or (((communit* or commune or communes or village* or local* or rural or suburban or urban or district* or quarter* or slum* or family or primary or citizen*) adj3 (health* or medicine or nurs*)) or (health* adj2 promot*) or (health* adj2 service*) or (health adj2 (center* or centre*)) or health planning* or health engagement* or (healthcare adj2 deliver*) or health disparit* or ((social or digital*) adj isolat*) or (communit* adj3 build*) or resilien* or ((social or mental) adj (wellbeing or well-being)) or ((migrant* or immigrant* or refugee* or asylum seek* or minorit* or vulnerable*) adj3 health*) or violence).ab,ti,id.)) not (editorial or correspondence or conference).pt.

##### ERIC Educational Resources Information Center Database (via OvidSP)

(Participatory Research/ or Action Research/ or Citizen Participation/ or Community Involvement/ or community action/ or Participative Decision Making/ or Social Networks/ or ((participat* adj3 (research or approach* or assistance or design* or method* or framework* or model or models)) or (partner* adj3 approach*) or “action research” or “community research” or “community based research” or (community-partnered adj3 research) or ((communit* or citizen* or residen* or minorit* or marginali* or rural or urban or suburb*) adj3 (engag* or empower* or attachment* or participat* or involv* or implement* or action* or interact* or activ* or collaborat* or partner* or respon* or work or worked or working)) or “civic engagement” or “participatory spaces” or “social mobilization” or “social mobilisation” or ((mobiliz* or mobilis*) adj3 (work* or CHW*)) or “co-creat*” or cocreat* or “co-prod*” or coprod* or “co-design*” or codesign* or “collaborat* creation” or co-construct* or coconstruct* or “collaborat* product*” or “collaborat* design*” or “community creation” or “community product*” or “community design*” or CBPR or caremongering).ab,ti.) and (COVID-19/ or pandemics/ or (coronavir* or corona virus* or HCoV* or nCoV* or 2019nCoV or 2019-novel CoV or covid*2 or covid-19 or covid19 or sars-cov* or sarscov* or Sars-coronavirus* or “corona pandemi*” or (pandemic* adj3 respon*) or “corona cris*” or Severe Acute Respiratory Syndrome Coronavirus*).mp.) and (community health services/ or mental health/ or rural areas/ or rural population/ or urban areas/ or urban population/ or slums/ or minority groups/ or disease control/ or “resilience (psychology)”/ or social isolation/ or health promotion/ or (((communit* or commune or communes or village* or local* or rural or suburban or urban or district* or quarter* or slum* or family or primary or citizen*) adj3 (health* or medicine or nurs*)) or (health* adj2 promot*) or (health* adj2 service*) or (health adj2 (center* or centre*)) or health planning* or health engagement* or (healthcare adj2 deliver*) or health disparit* or ((social or digital*) adj isolat*) or (communit* adj3 build*) or resilien* or ((social or mental) adj (wellbeing or well-being)) or ((migrant* or immigrant* or refugee* or asylum seek* or minorit* or vulnerable*) adj3 health*) or violence).ab,ti.)

##### CINAHL (EBSCOhost)

Cumulative Index to Nursing and Allied Health Literature CINAHL with Full Text (via EBSCOhost) Search Options: Expanders - Apply equivalent subjects, Search modes - Find all my search terms (MH “Action Research” OR MH “Consumer Participation” OR MH “Citizen Science” OR MH “Community Networks” OR ((participat* N3 (research OR approach* OR design* OR method* OR framework* OR model OR models)) OR (partner* N3 approach*) OR “action research” OR “action science” OR “community research” OR “community based research” OR (community-partnered N2 research) OR ((civic OR communit* or citizen* OR residen* OR minorit* OR marginali* OR rural OR urban OR suburb*) N2 (engag* OR empower* OR attachment* OR participat* OR involv* OR implement* OR action* OR interact* OR activ* OR collaborat* OR partner* OR respon* OR work OR worked OR working)) OR “participatory spaces” OR “social mobilization” OR “social mobilisation” OR ((mobiliz* OR mobilis*) N2 (work* OR CHW*)) OR “co-creat*” OR cocreat* OR “co-prod*” OR coprod* OR “co-design*” OR codesign* OR “collaborat* creation” OR co-construct* OR coconstruct* OR “collaborat* product*” OR “collaborat* design*” OR “community creation” OR “community product*” OR “community design*” OR CBPR OR caremongering)) AND (MH “Coronavirus” OR MH “Coronavirus Infections” OR MH “COVID-19” OR MH “COVID-19 Pandemic” OR MH “SARS-CoV-2” OR (coronavir* or “corona virus*” or HCoV* or nCoV* or 2019nCoV or “2019-novel CoV” or covid or covid-19 or covid19 or sars-cov* or sarscov* or Sars-coronavirus* or “corona pandemi*” or “corona cris*” or “Severe Acute Respiratory Syndrome Coronavirus*”)) AND (MH “Health Services Research” OR MH “Infection Control+” OR MH “Community Health Services+” OR MH “Community Mental Health Services” OR MH “Community Mental Health Nursing” OR MH “Community Health Nursing” OR MH “Community Health Workers” OR MH “Community Health Centers” MH “Rural Health Services” OR MH “Urban Health Services” OR MH “Suburban Health” OR MH “Urban Health” OR MH “Health Care Delivery” OR MH “Health Status Disparities” OR MH “Sexual and Gender Minorities” OR MH “Hardiness” OR MH “Social Isolation” OR MH “Health Promotion” OR (((communit* OR commune OR communes OR village* OR local* OR rural OR suburban OR urban OR district* OR quarter* OR slum* OR family OR primary OR citizen*) N2 (health* OR medicine OR nurs*)) OR (health* N2 promot*) OR (health* N2 service*) OR (health N2 (center* OR centre*)) OR health planning* OR health engagement* OR (healthcare N2 deliver*) OR health disparit* OR ((equal* OR inequal* OR equit* OR inequit*) N1 (access*)) OR ((social OR digital*) N1 isolat*) OR (communit* N2 build*) OR resilien* OR ((social OR mental) N1 (wellbeing OR well-being)) OR ((migrant* OR immigrant* OR refugee* OR asylum seek* OR minorit* OR vulnerable*) N2 (health*)) OR violence)) NOT (PT commentary OR PT editorial OR PT letter)

#### **ClinicalTrials.gov** https://www.clinicaltrials.gov/ (via Expert Search)

((“participatory research” OR “action research” OR “community research” OR “community based research” OR “civic engagement” OR “community engagement” OR “public engagement” OR “citizens engagement” OR “co-production” OR “co-design” OR “co-creation” OR partnership OR collaboration) AND (coronavirus OR “corona virus” OR HCoV OR ncov OR covid OR “covid 19” OR “sars cov” OR “sars-cov” OR “sars coronavirus” OR “Severe Acute Respiratory Syndrome Coronavirus”))

**Google Scholar** (first 200 results, out of 17’100 results – sorted by relevance) participatory|“action research”|“citizen science”|“civic|citizen|community|resident engagement”|“co creation|production|design|construction”|CBPR coronavirus|“corona virus|pandemic|crisis”|HCoV|ncov|covid “community health”|well-being|wellbeing

